# Developing services for long Covid: lessons from a study of wounded healers

**DOI:** 10.1101/2020.11.13.20231555

**Authors:** Emma Ladds, Alex Rushforth, Sietse Wieringa, Sharon Taylor, Clare Rayner, Laiba Husain, Trisha Greenhalgh

**Affiliations:** Nuffield Department of Primary Care Health Sciences, University of Oxford; Central and North West London NHS Foundation Trust, Imperial College London, United Kingdom; Division of Psychiatry, Imperial College London, United Kingdom; Independent Occupational Physician, Manchester

**Author notes:** **Corresponding author:** E. Ladds, Nuffield Department of Primary Care Health Sciences, University of Oxford, Oxford OX2 6GG, UK.

**Keywords:** Post-acute Covid-19, Chronic Covid-19, long Covid, qualitative study, new service model, quality standards

## Abstract

Persistent symptoms lasting longer than 3 weeks are thought to affect 10-20% of patients following Covid 19 infection. No formal guidelines exist in the United Kingdom for treating ‘long Covid’ patients and services are sporadic and variable, although additional funding is promised for their development.

In this study narrative interviews and focus groups are used to explore the lived experience of 43 healthcare professionals with long Covid. These individuals see the healthcare system from both professional and patient perspectives thus represent an important wealth of expertise to inform service design.

We present a set of co-designed quality standards highlighting equity and ease of access, minimal patient care burden, clinical responsibility, a multidisciplinary and evidence-based approach, and patient involvement and apply these to propose a potential care pathway model that could be adapted and translated to improve care of long Covid patients.

**Summary box:** *What is known?:*

▪ Persistent symptoms (“long Covid”) occur after Covid-19 in 10-20% of sufferers
▪ Services to manage and rehabilitate patients with long Covid are not yet optimal
▪ UK healthcare workers experience at least a threefold greater risk of Covid-19 infection and face significant occupational exposure
▪ Healthcare workers with long Covid can offer important insights into service design and development

*What is the question?:*

▪ What are the experiences of healthcare workers with long Covid and what are the implications from these for service development?

*What was found:*

▪ Healthcare workers experienced a confusing novel condition that imposed high levels of uncertainty and a significant personal and professional impact.
▪ Using professional contacts, patient- and professional Mindlines, support groups and Communities of Practice all helped to minimize this uncertainty and high quality therapeutic relationships were essential to cope with it.
▪ Many experienced a lack of compassion during interactions with the healthcare system and were frustrated by challenges accessing, or absence of, appropriate services.
▪ Suggestions for improvement included an integrated, multi-disciplinary assessment and rehabilitation service; a set of clinical quality standards; and co-created research and service development.

*What is the implication for practice now?:*

▪ This study supports and extends the principles outlined in recently-developed NHS long Covid quality standards and will inform and support design of dedicated long Covid services.

## Introduction

In the UK, Covid-19 has struck a healthcare system already at breaking point. The number of GPs per 100,000 is at its lowest since 2003,^1^ there is a nursing shortage of 40,000,^2^ and 40.3% of NHS workers reported feeling unwell as result of work related stress in the last year.^3^ For healthcare workers, the moral injury of caring for patients in the context of a near-incurable virus alongside personal danger, fear of placing loved ones at risk, extended shifts, disrupted processes, rota gaps, and wider social restrictions have only compounded pressures.^4^

Studies suggest that 10-20% of symptomatic individuals with Covid19 suffer persistent symptoms, with 1-3% affected at 12 weeks. This long Covid syndrome manifests in a relapsing-remitting range of multi-system symptoms and affects all individuals including those who were previously fit and healthy.^5^ Preliminary guidance exists to support management^6^ and the National Institute for Health and Clinical Excellence, Scottish Intercollegiate Guidelines Network, and Royal College of General Practitioners are collaborating on a definitive guideline. However, whilst NHS England has allocated funding for a new Long Covid service,^7^ the detail remains unconfirmed.

Healthcare workers with long Covid are ideally placed to contribute to organisational sensemaking required to respond to a new disease and become partners in service development. Alongside the unique perspective of being both within (“emic”) and external (“etic”) to the system,^8^ many have also taken on key advocate and leadership roles within the long Covid community.^9^ In this study, we sought to explore the experiences of healthcare workers with long Covid to develop a set of quality standards and potential care pathway model for management of long Covid.

## Methods

Details of study set-up, governance and methods are published elsewhere.^10^ The study was conducted between May and September 2020 and received ethical approval from the East Midlands – Leicester Central Research Ethics Committee (IRAS Project ID: 283196; REC ref 20/EM0128). The dataset comprised 114 participants who contributed through online focus groups, individual narrative interviews, or symptom diaries and statements. This analysis focused on 43 respondents. Individuals were recruited through online long Covid support groups and social media, followed by a snowballing strategy and consented by email or verbally. Participants were invited to tell their stories uninterrupted. Audio and video recordings and contemporaneous notes were transcribed in full, deidentified and entered onto NVIVO software version 12. Text were grouped into broad categories and subsequently refined using the theoretical lenses described in the discussion. Two clinically qualified individuals with long Covid, listed as co-authors, assisted with data interpretation. All participants were sent a summary and invitations were encouraged to correct any errors or misinterpretations.

## Results

### Description of dataset

Participant demographic details are shown in Table 1. Despite our efforts to balance for gender and ethnicity, the final sample was skewed to 81% female and 84% White – in line with the gender representation in long Covid support groups but less ethnically diverse than the UK population.^11^ In total around 500 pages of transcripts were produced. Our broad coding produced six over-arching themes: uncertainty, use of Mindlines, support groups and Communities of Practice, therapeutic relationships and roles, professional identity and practice, and suggestions for service improvement.

**Table□1:**
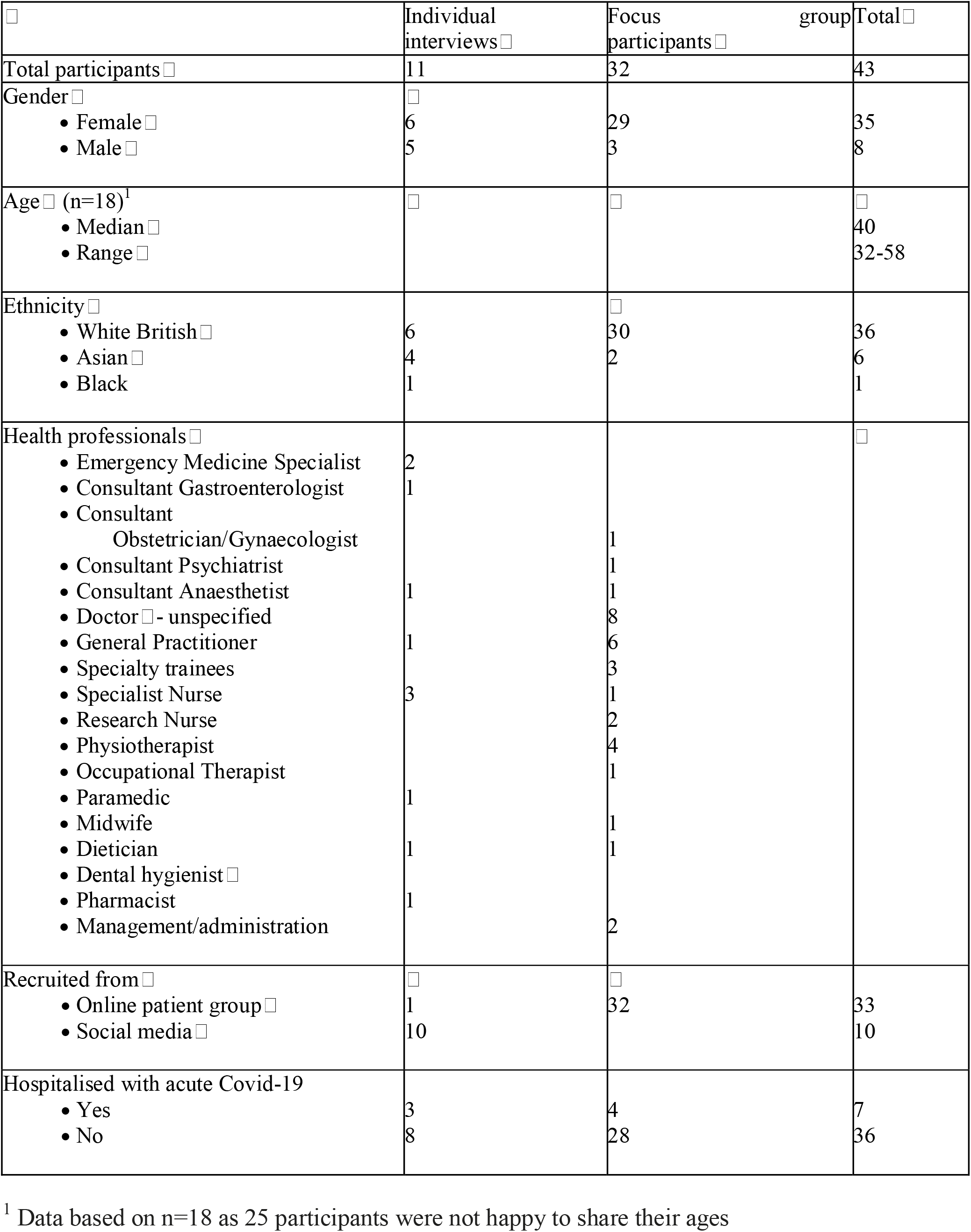
Participant characteristics□.

### Uncertainty

Participants reported a constellation of persisting, atypical symptoms that did not fit an expected pattern based on their pre-existing professional knowledge. Both participants and healthcare professionals caring for them found this challenging and attempted to mitigate the uncertainty by placing symptoms within a framework of pathologies or conditions they *did* understand i.e. negotiating referrals for familiar investigations or specialist reviews. However, many recognized the limitations of this approach, emphasizing the need for further research and knowledge development.

> She went and did a Computed Tomography Pulmonary Angiogram and I said what are we doing with these tests because I had the tachycardia as well and there was flattening all over my ECG and she said well we don’t know what to do with long Covid what we’re doing at the moment is what we always do in these situations we’re ruling out other things but if the answer ends up being long Covid we don’t know. […] I just guard a bit against this. We have the tests and reassurance but we’ve still got crushing central chest pain, we still get breathless eating soup, we’re no better for the investigations, the investigations aren’t a treatment…
>
> (Participant from doctors FG1)

### Use of Mindlines

The absence of evidence frequently turned participants towards other sufferers, professional contacts, and online communities to make sense of their experience and navigate the best way forwards without formalized protocols. Alongside practical benefits such as the use of contacts or colleagues to navigate the system and access investigations or specialist reviews, a deeper generation of ‘Mindlines’ emerged i.e. ‘internalised and collectively reinforced tacit guidelines’, which are frequently used by professionals in preference to formal guidance.^12^

> I was fortunate enough to have colleagues working in the hospitals local to me [hospital names]. Basically as they were trying to put in central lines and realising blood was like glue they would be ringing me up and saying have you thought about doing this, that and the other as we started to understand the full plethora of systems that were affected by the virus.
>
> (Individual interview AP)

Alongside professional contacts, many participants reported the support offered by patient-developed Mindlines developed from the experiences of unwell colleagues or through social media communities. This was particularly relevant where experiences ran counter to ‘official’ guidance. For example, many participants emphasized the physical and psychological impact of the lag before persistent symptoms were incorporated in a wider folk understanding of long Covid, in the face of government advice detailing an expected time course of 2-3 weeks.

> At week 7 I was offered an antibody test at work, which came back negative. That day was really bad. I thought I was going mad - I didn’t know of anyone else who was taking this long to get better and I couldn’t understand why. Did I have something else? Unlikely for an ED consultant during a pandemic, but not impossible. Why didn’t I have antibodies, would anyone believe that I had COVID? A few days later colleagues and articles on social media started to describe people with symptoms like mine and I began to feel like I wasn’t alone.
>
> (Individual Interview RM)

### Support Groups and Communities of Practice

Participants frequently expressed how helpful they found the Long covid social media support groups. These offered a means of sharing health-related information and enabled the development of patient – and professional – Mindlines. Alongside these practical benefits, groups offered a sense of shared identity and belonging, enabling retrospection of experiences, development of narratives and in many ways becoming ‘Communities of Practice’.^13^

> I was struggling with work as well because one of my colleagues had said well isn’t Post Covid just anxiety and I think after my GP had that conversation she actually suggested that I set up a Facebook group for other doctors and then XX set one up that weekend and that’s kind of how that happened and it’s been really helpful; kind of shared experiences and realising that you’re not alone and kind of being a safe space.
>
> (Participant from doctors FG2)

### Therapeutic relationships and roles

Similar to online communities, positive interactions with healthcare workers were perceived as those that enabled listening and sharing of experiences with an honest acknowledgement of the uncertainties, acceptance of the evidence gap, and openness to draw on other resources – eg: colleagues or patient experience.

> I think if someone can acknowledge uncertainty then I think that really helps because I think we all know that nobody knows what to do with us but I think where it can become frightening is if they’re kind of claiming unwarranted certainty. So I think actually just saying well actually we don’t really know what’s going on but yes stick with us we’ll try and work it out.
>
> (Participant from doctors FG2)

Bearing witness to and sharing suffering – not necessarily ‘fixing’ it – was hugely significant. Participants had mixed feelings about the appropriateness of remote consultations in doing this. Whilst recognizing their necessity in the pandemic context, many emphasized the value and reassurance of face-to-face assessment, particularly given the disease’s impact on their ability to make clinical judgments and communicate this remotely.

> I’ve not actually physically seen my GP face to face on any of these occasions […] when I phone up it’s very much what are you thinking it might be, which when you’re not feeling well it’s very difficult. And then I always come off the phone quite dissatisfied. I’ve actually spent this morning in A&E because I thought I’ve never had a positive diagnosis of Covid. I’ve been unwell for what ten weeks now, I need to make sure it’s nothing else and I’ve not actually been examined at any point.
>
> (Participant from doctors FG1)

Clinician attitudes also impacted encounters, with confirmatory or personal biases resulting in clinician denial. This compounded the uncertainty participants felt governed many GPs, particularly when faced with well-read, expert patients, occasionally resulting in a dilemma of authority that impeded the quality of therapeutic relationships and outcomes.

> My last interaction with my GP was in June. I asked about my lungs, and he said what do you want me to do about it? You tell me. I have no idea. It felt very dismissive and it felt like I know GPs don’t want patients to look up things on the internet and come and say this is what I think’s wrong with me because in the normal world that’s definitely not what you want but that’s what every person with Covid’s been driven to because the response from GPs is I don’t know what to do. Nothing’s got any evidence so yeah sorry I can’t help. I went back to work after five weeks still very unwell because nobody believed in Long Covid in May, they just didn’t believe it.
>
> (Participant in Allied Health Professionals FG3)

Some participants found attempts at ‘shared decision making’ in the acute illness challenging, simply preferring the ‘patient’ role, whilst others welcomed this transparency. However, the clinical uncertainties later experienced around persistent symptoms and unknown prognosis were frightening for many, regardless of which ‘role’ was adopted and especially if compounded by a lack of therapeutic continuity or follow-up.

### Professional Identity and Practice

The uncertainty around symptom duration and personal impact of the disease compounded professional tensions. Participants emphasized their strong work ethic and teamworking role and feared colleagues would perceive them as ‘shirkers’. However, individuals also had good insight into their physical and mental limitations and expressed fear of coming back to work too soon risking reputational damage or medico-legal ramifications.

> The medical legal aspect is huge and I think certainly feels that way as a GP and it’s scary to not be able to recognise potentially where you have deficits because if you can’t recognise them then that’s an unknown unknown in what can you do with that.
>
> (Participant in doctors FG1)

There was also much anger and frustration, particularly around accessing care or responses they had received from an unkind, uncompassionate ‘system’. This was particularly salient when experienced from ‘the other side’ as a patient.

> I know quite a few of the people at the health centre personally and I think they’re all really good people and really caring people but the experience from my end has not been one of care particularly.
>
> (Participant from Allied Health Professional FG3)

### Suggestions for service improvement

Participants emphasized the duty they felt as eloquent, informed ‘expert patients’ with knowledge of ‘the system’ and personal experience of its limitations to advocate for service improvements.

> I mean not, not to sort of self-grandiose our group but there’s a certain responsibility to put down our experiences so they can be opened up to other people who don’t have the language and the access that we potentially have to communicate it to primary health care to access the services that need to be put in place for them.
>
> (Participant from doctors FG2)

Difficulty accessing investigations for persisting symptoms and the siloed, single-organ, and unintegrated nature of the system led participants to propose holistic, ‘one-stop-shop’ services integrating multiple specialties. A co-designed potential structure is presented in figure 1.

**Figure 1.**
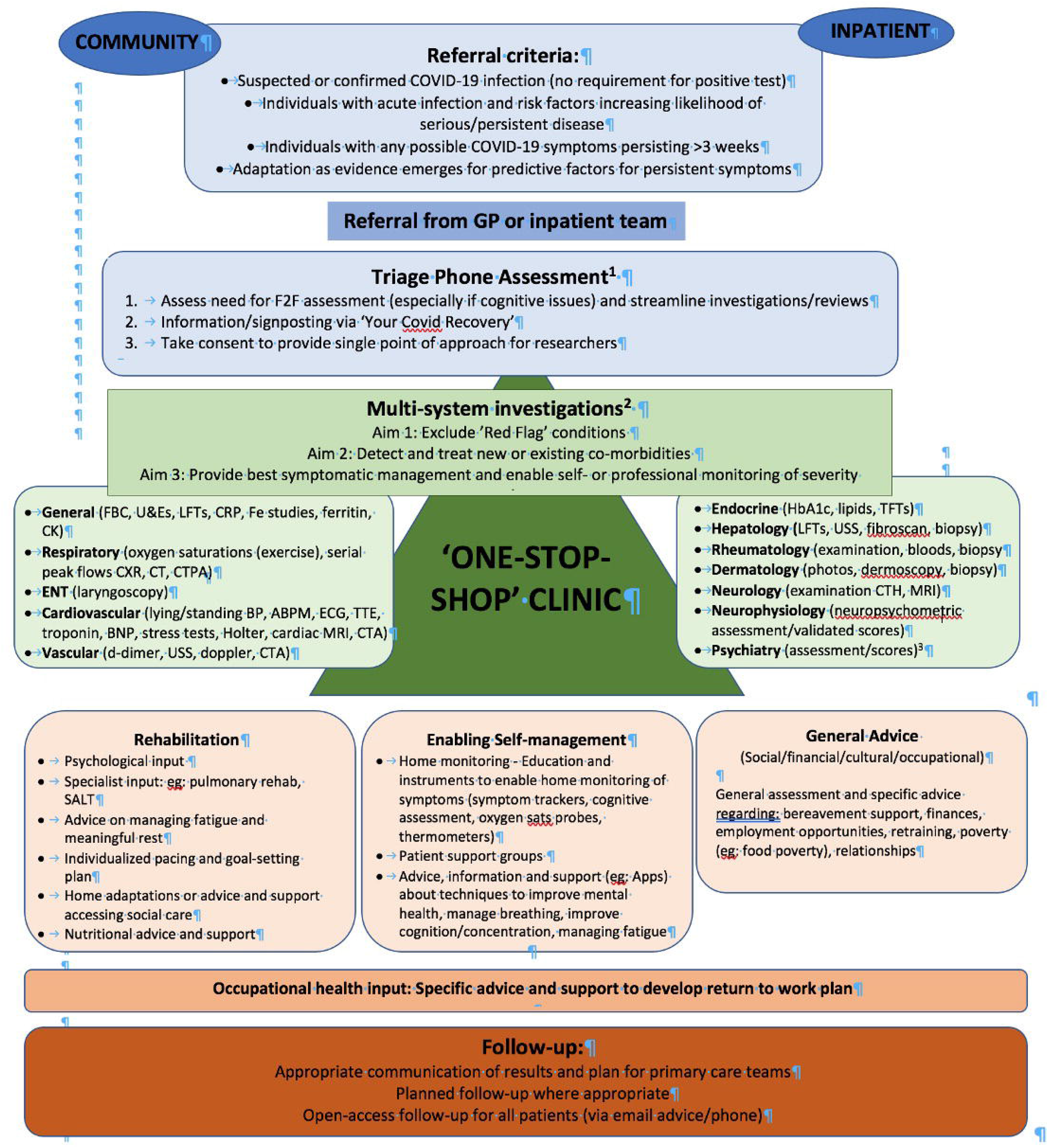
^*1*.^ *For example using the COVID 19 Yorkshire Screening Tool or post-COVID-19 Functional Status Tool (*https://www.england.nhs.uk/coronavirus/wpcontent/uploads/sites/52/2020/10/C0840_PostCOVID_assessment_clinic_guidance_5_Nov_2020.pdf - Appendices C and D) ^*2*.^ Possible specialist investigations – not a definitive list ^*3*.^ *FBC – Full Blood Count; U&Es – Urea and Electrolytes; LFTs – Liver Function Tests; CRP – C-Reactive Protein; Fe studies – Iron studies; CK – Creatinine kinase; CXR – Chest Radiograph; CT – Computed Tomography Scan; CTPA – Computed Tomography Pulmonary Angiogram; BP – Blood Pressure; ABPM – Ambulatory Blood Pressure Monitoring; ECG - Electrocardiogram; TTE – Transthoracic Echocardiogram; BNP – Brain-Natriuretic Peptide; MRI – Magnetic Resonance Imaging (study); CTA – Computed Tomography Angiography; USS – Ultrasound Scan; TFTs - Thyroid Function Tests; CTH – Computed Tomography (scan) of the Head*

Such services should be designed to exclude red flag conditions, detect and treat co-morbidities or complications, and support symptomatic management. They must be multidisciplinary, run by generalists rather than a single specialty and ideally be situated in primary care to enable continuity in follow-up and rehabilitation, with appropriate links for secondary care referral. However, it was recognized they must not fall on GPs and must be appropriately resourced.

> My expectation of such a clinic would be to rule out treatable causes or complications, based on our symptoms. And then active involvement with physiotherapies and occupational therapies maybe a psychologist […] we now know that Covid is a multi-system disease so the fact that you don’t display signs of respiratory infection doesn’t mean that you don’t have a problem.
>
> (Participant Allied Health Professionals FG1)

The importance of consistent, equitable, and inclusive access criteria was clear – specifically with no prerequisite for a positive PCR or antibody Covid19 test result, merely referral from a healthcare professional. Linked to this, participants advocated further training of GPs, raising the possibility of a long Covid practice lead. Integration and education of allied health professionals, psychologists and occupational health teams were also emphasized, given the acknowledgement that ‘treatment’ would frequently be time and personalised rehabilitation depending on deficits and organ complications in the context of a caring therapeutic relationship.

Reflexivity and improvement, sharing of knowledge and experiences, and an encouragement of further research were also essential. Despite these ideals, participants remained sceptical about the likely translation of their proposals due to financial restraints, personnel shortages, and the nature of change within the NHS.

> I don’t know how anyone is gonna create this sort of a clinic it just seems, I mean we know how the NHS works and it just seems like a far flung reality
>
> (Participant from doctors FG2)

## Discussion

### Summary of key findings

This qualitative study of 43 UK healthcare workers with long Covid highlights the great uncertainty experienced by patients and healthcare professionals around a novel condition, compounded by pre-existing confirmatory biases and an absence of guidelines or care pathways. Applying existing disease frameworks, utilizing professional networks, and the development of professional and patient Mindlines and Communities of Practice were all attempts to minimize this uncertainty, whilst therapeutic relationships providing supportive continuity and follow-up best enabled individuals to cope with the tension of the unknown.^14^

The personal and professional challenges of long Covid were exacerbated by frustrations and callousness participants experienced in their encounters with the healthcare system. Given their status of both professionals and patients they felt a responsibility to aid service improvements and advocated for holistic, multidisciplinary and multisystem services and research initiatives.

### Comparison with theoretical literature

Sociological theories of the clinician as patient abound. Hahn and Klitzman highlighted the concept of the ‘medical self’ where clinicians behave like a doctor even when being a patient. ^15,16^ This is borne out within our data and compounds and exacerbates the uncertainty. Role ambiguity, reversal, or vacillation along a spectrum between the doctor’s and patient’s roles heightens anxiety^17^ and may heighten a clinician’s personal sense of frailty, creating a tension which could result in minimization of a patient’s issues as a protective coping mechanism.

‘Wounded healers’ i.e. professionals with experience of patienthood are frequently said to have developed deeper understanding and wisdom. In ethnomedicine, healers are often chosen *because* they have experienced severe illnesses^15^. In this case, despite exacerbating role confusion, ‘medical patients’ felt a duty to advocate for future patients^18^ through experience-based [co-]design (EBCD) of long Covid services. The duality of their ‘emic’ and ‘etic’ selves with dual vantage points of the healthcare system situated them particularly well to do so.^8^

Experience-based [co-]design was developed to ensure health services were designed around the patient experience.^19^ Grounded in the perceptions and reactions of individual patients this collective sensemaking process produces new understandings that can be pragmatically applied to frontline services^19^ – a process enjoyed by participants in this study through participation in focus groups; online communities and authorship groups advocating further action.^9^

### Implications for services

Alongside a potential multidisciplinary care pathway structure (figure 1), this study provides supportive evidence for a set of quality principles: ‘Long Covid ABCDE’ (box 1),^10^ many of which accord strongly with a manifesto written by a group of doctors with long Covid.^9^ As well as minimizing patient suffering, it is hoped implementation would reduce mortality, improve patient outcomes, and optimize return to employment.

#### Box 1

**‘ABCDEF’ of Long Covid Clinical Quality Standards**

1. **ACCESS:** Services should be accessible to everyone with long Covid regardless of Covid positive PCR, antibody test, or hospitalisation status. However, as the evidence base and diagnostic criteria develop, referral criteria to specialist services may need refinement to ensure optimal resource utilisation.
2. **BURDEN OF ILLNESS:** The burden of treatment needs to be minimized. A “one-stop shop” model could facilitate this such as those developed for other complex conditions eg: memory impairment. Alternatively, a shared care model, with clearly defined care pathways disseminated between the community and secondary care is likely to be more achievable for many localities and accords with the Department of Health’s long-term conditions model. Such approaches have been applied for over 10 years to multisystem conditions including inflammatory arthritides and diabetes or mental health problems. Outcomes are non-inferior to those achieved through secondary care management^26^ and may benefit from enhancing patients’ self-efficacy, confidence and satisfaction, and cost-efficacy.^27^ Successful integration relies on coordination between commissioners and providers, adequate financial resources and appropriate governance structures.
3. **CLINICAL RESPONSIBILITY AND CONTINUITY OF CARE** A named clinician must be responsible for the patient at every stage. Management and relationship continuity combined with clinical responsibility are essential in any multisystem or chronic condition and are associated with patient and physician satisfaction, reduced clinician collusion of anonymity, physician trust and therefore treatment adherence, problem recognition and quality of management, and reduced healthcare utilisation and associated costs.^28^ Such continuity has also been shown to improve outcomes in rehabilitation care settings - of particular relevance for long Covid.^29^
4. **MULTI-DISCIPLINARY REHABILITATION SERVICES** Services must be multi-disciplinary. Integrated care requires professionals and practitioners from different backgrounds to work together to improve patient outcomes. Multi-disciplinary team structures provide a shared identity and purpose encouraging trust, a more holistic and person-centred practice, minimizing errors and associated harms, improving efficiency of resource utilisation and reducing professional isolation and stress.^30^ From cancer care to rehabilitation, such approaches have been widely implemented within the NHS to improve clinical outcomes, patient and family experience and professional satisfaction.
5. **EVIDENCE-BASED STANDARDS** Evidence-based standards must be developed to guide consistent symptom management. These should guide the exclusion of potentially serious complications, management of new or existing co-morbidities, and holistically address symptoms and should acknowledge the best available evidence or consensus of best practice in the context of a rapid, dynamic context, rather than holding out for randomized controlled trials.
6. **FURTHER DEVELOPMENT OF THE KNOWLEDGE BASE AND CLINICAL SERVICES** Patients must be offered the opportunity for involvement in further research and service development, which should be enabled by the development of registries of patient data and facilitated through long Covid services. This reflects the key principles of patient-centred medicine and such experience-based co-design also improves alignment of research and service aims with patient priorities; development of meaningful and useful studies and services, in which patients are keen to participate; identification and minimization of barriers to access or utilization; enhanced patient education, information dissemination, and application of research findings; improved patient experience and cost efficiency.

### Strengths and limitations of the study

To our knowledge, this is the largest and most in-depth qualitative study of healthcare workers with long Covid published in the academic literature with a focus on the under-researched majority who were never hospitalized. However, the population sample was drawn exclusively from the UK and may have inadequately captured the perspectives of ethnic minority groups. Further strengths and limitations are discussed elsewhere.^10^

### Comparison with previous empirical studies

Our findings affirm the experience of healthcare professionals with long Covid published in narrative reviews,^20^ commentaries,^21^ and manifestos.^9^ Like our own dataset, these emphasise the disorienting uncertainties of this new illness; the stigma of not being believed; frustration of scientific and medical ignorance; and problems accessing services.

‘Wounded healers’ are extensively represented throughout the illness experience literature. Our population did not reflect the ethos of invulnerability reported in other narrative accounts,^22^ however the challenges of the patient role, importance of therapeutic relationships and compassionate care, inhumanity of ‘the system’ and use of professional status to manipulate it, workforce culture and guilt that promote return to work, and impact on professional understanding and practice all emerged here.^23^ Interestingly, as with long Covid, many wounded healers describe their experiences in non-academic forums such as blogs, memoirs, or media outlets, particularly in times of illness that challenges or changes life or identity eg: burnout,^24^ or palliative care,^25^ reflecting a need to attempt a narrative sense-and meaning-making process.

## Conclusion

This study of healthcare professionals with long Covid provides further evidence of approaches to minimize and live with the great uncertainty surrounding the condition, capitalizing on their dual status as professionals and patients to support quality standards and suggest improvements for long Covid services within the NHS.

## Data Availability

All data is available within the manuscript.

